# Prediction of Heart Failure based on Multimodal Data from MIMIC-IV

**DOI:** 10.64898/2026.08.17.26360588

**Authors:** Anna Lutz, Fabio Hellmann, Elisabeth André

## Abstract

Heart failure (HF) affects over 64 million people worldwide and remains a leading cause of cardiovascular mortality. Early identification of patients at risk is essential for timely treatment and to support hospital and primary care physicians. This study compares XGBoost and a Transformer-based bidirectional cross-attention model using multimodal data to assess whether deep learning (DL) approaches can outperform classical machine learning (ML) methods for early HF prediction. We identified HF and non-HF patients from the MIMIC-IV database using ICD-9/10 codes, supplemented by clinical evidence from laboratory results, radiology, and discharge notes. Furthermore, we defined a 48-hour prediction window prior to the first clinical evidence of HF. Structured features were engineered from rolling-window statistics and clinical thresholds. Both XGBoost and Transformer models were trained on multimodal data and compared through an ablation study. Finally, we developed a dashboard using a small set of laboratory and medication features to deliver a 48-hour HF risk estimate, aiding clinician diagnosis. Multimodal models outperformed single-modality models across both architectures. The multimodal XGBoost model achieved the highest performance (F1 of 0. 8773 and PR-AUC of 0.9402), while the multimodal Transformer achieved slightly lower performance (F1 0.8635, PR-AUC 0.9209). Structured data contributed most to XGBoost (PR-AUC of 0.9163), whereas clinical notes were better captured by the Transformer (PR-AUC of 0.8220). Explainable dashboards further enhance transparency and usability by delivering quantitative 48-hour risk estimates from minimal features. This demonstrates, in this setting, that traditional ML can outperform DL models such as Transformers on tabular-dominated, multimodal clinical prediction tasks while preserving interpretability, underscoring decision-support systems’ potential to aid timely diagnosis.

https://doi.org/10.1515/cdbme-1

## 1 Introduction

Heart failure (HF) is a complex syndrome and is expected to be the third leading cause of cardiovascular death in Germany in 2024, accounting for 11.09% of cases [3]. It results from functional or structural heart changes that reduce pumping efficiency and increase cardiac pressure, manifesting as fatigue, ankle swelling, and edema [10]. HF has high clinical and economic relevance. In North Rhine-Westphalia, HF was the most common reason for outpatient hospital stays in 2024 [5]. In Germany overall, it generated treatment costs of 9.241 million euros in 2023 [2]. Despite advances in treatment, survival remains limited according to the Rotterdam Study, with 11% of patients dying within one year and 41% within five years after diagnosis [12]. Early detection is challenging due to a gradual onset and nonspecific symptoms [11] but allows outpatient stabilization and medication optimization. Because HF diagnosis relies on a combination of laboratory values, clinical history, and physician observations, large multimodal intensive care datasets provide a suitable basis for developing predictive models. The public MIMIC-IV dataset contains over 220,000 ICU admissions with structured clinical data and unstructured text from radiology and discharge reports [7], making it particularly relevant for multimodal HF prediction. Previous research has compared XGBoost, a machine learning (ML) approach, with Transformer-based deep learning (DL) models on structured single-modality data [8]. Building on this, this study evaluates both methods in a multimodal setting combining structured and unstructured data and integrates the models into a dashboard to support diagnosis with minimal input requirements.

## 2 Methods

This section describes the datasets, cohort structure, and model training and testing. Figure 1 illustrates the individual steps. The conducted research is not related to either human or animals use.

**Figure 1.**
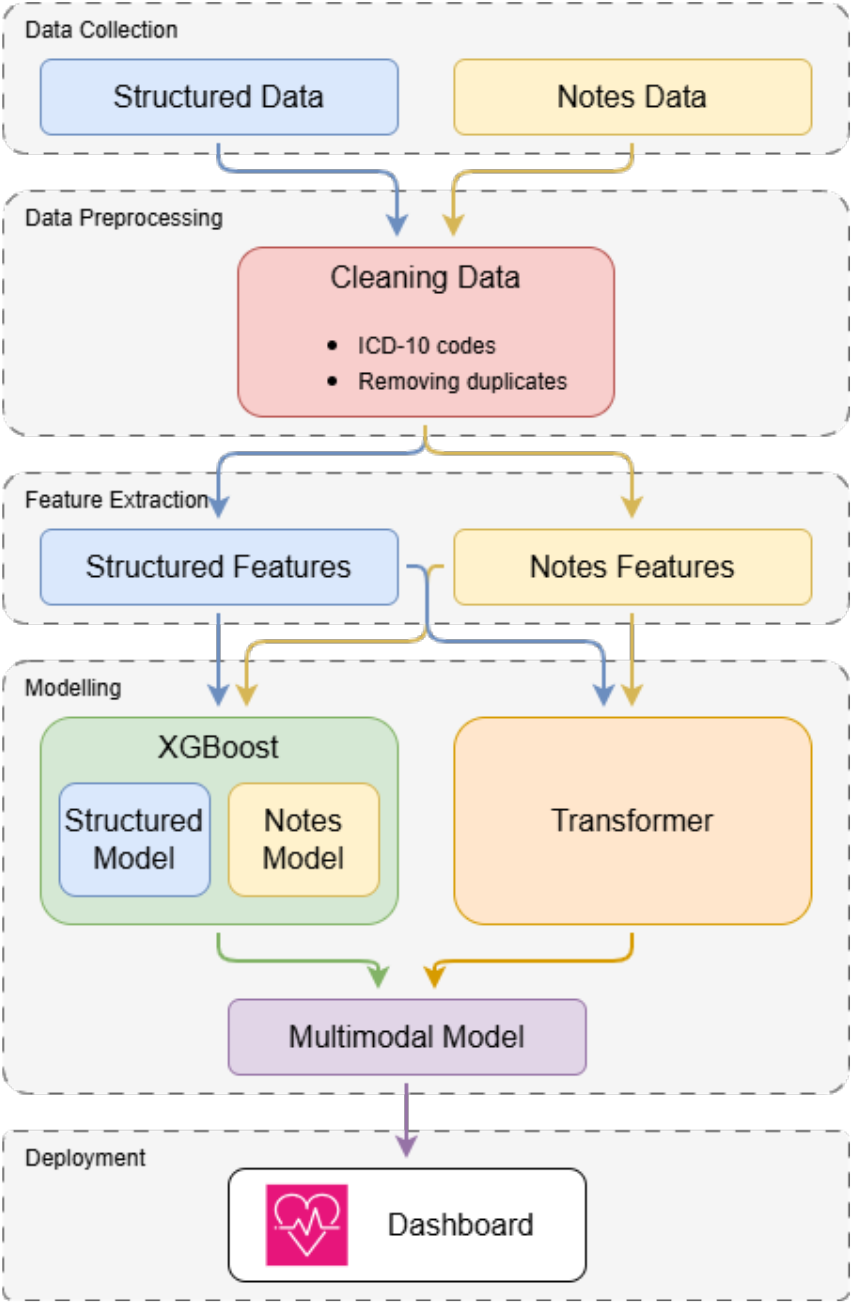
Pipeline of the study. Yellow areas indicate notes, blue represents structured data, and purple shows multimodal data. Both Transformer and XGBoost were trained on all three data types, with the multimodal model integrated into the dashboard.

### 2.1 Dataset and Cohort Construction

For this study, the MIMIC-IV dataset (version 3.1) [7] and the MIMIC-IV-Note dataset (version 2.2) [6] were used. HF patients were identified via ICD-9 and ICD-10 codes. Patients without reliably determined HF onset beyond ICD diagnosis were excluded. Non-HF controls were matched to HF patients by age, sex, and ICU status using stratified random sampling to ensure comparable distributions of these covariates while preserving cohort size and statistical power. The final cohort included 68,507 HF and 53,096 non-HF patients, with similar age and sex distributions, approximately 47% female and 52% male in both groups.

### 2.2 Prediction Window

The 48-hour HF prediction window was defined using the earliest timestamp from four sources, including mentions in clinical notes, Brain Natriuretic Peptide (BNP) values of at least 400 pg per mL, first administration of loop diuretics, and signs of pulmonary edema or vascular congestion in radiology reports [10]. Non-HF controls received pseudo-onset times.

### 2.3 Feature Engineering

The 48-hour forecast window was split into eight six-hour intervals. For each clinical feature, such as laboratory measurements and medication doses, statistics such as the mean, standard deviation, minimum, and maximum were computed for each item within each bin. By computing statistics for each bin, this representation preserves the temporal dynamics of individual measurements. In addition, overall statistics were generated for each item across the entire window to capture cumulative trends. Unstructured data was preprocessed by removing stop words and converting text to lowercase. For XGBoost, all admission notes were combined into a single summary, whereas the Transformer retained notes in their original chronological order.

### 2.4 Model Architectures

Both algorithms were evaluated in unimodal and multimodal configurations. All models were trained and tested on the same dataset with patient-level splits of 64% for training, 16% for validation, and 20% for testing.

#### 2.4.1 XGBoost

The XGBoost model used features from feature engineering (Section 2.30). Unstructured notes were processed as TF-IDF vectors (40–15,000 tokens per patient). The multimodal model concatenates structured and unstructured inputs.

#### 2.4.2 Transformer

The Transformer-based bidirectional cross-attention model uses two parallel encoders. The structured encoder is a multi-layer perceptron with batch normalization producing a 256-dimensional embedding. The note encoder consists of three Transformer layers with four attention heads and aggregates the note sequence using attention pooling. The two modalities are fused through bidirectional cross-attention and further processed by a deep MLP with confidence weighting. Training was performed with AdamW over 15 epochs. Model performance was evaluated using bootstrap analysis with 1,000 repetitions to determine the 95% quantile. This resampling-based method provides estimates of bias, standard errors, and confidence intervals [4]

### 2.5 Dashboard

Both multimodal models were integrated into a clinical dashboard that provides patient-specific HF risk scores with feature explanations. Clinicians can estimate a patient’s 48-hour HF risk using key inputs such as vital sign and lab values, with the probabilities serving as a diagnostic aid.

## 3 Results

The following section presents the performance metrics for the models, summarized in Table 1. For unimodal models on structured data, XGBoost achieved a PR-AUC of 0.9369 and an F1 score of 0.8715, while the Transformer reached a PR-AUC of 0.9194 and an F1 of 0.8575. On the performance front, both models performed worse, but the Transformer outperformed XGBoost across most metrics, with PR-AUC of 0.8220 and F1 of 0.7663, compared to 0.7198 and 0.7071 for XGBoost. Multimodal integration improved performance, with XGBoost achieving the highest overall scores (F1 0.8773, PR-AUC 0.9437) and the multimodal Transformer achieving F1 0.8635 and PR-AUC 0.9209.

**Table 1.**
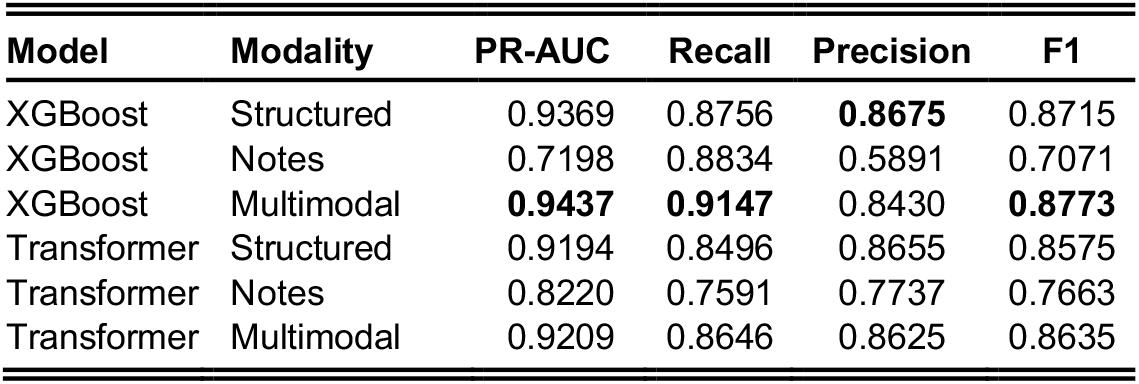
Performance metrics of XGBoost and Transformer models across different data modalities.

| Model | Modality | PR-AUC | Recall | Precision | F1 |
| --- | --- | --- | --- | --- | --- |
| XGBoost | Structured | 0.9369 | 0.8756 | **0.8675** | 0.8715 |
| XGBoost | Notes | 0.7198 | 0.8834 | 0.5891 | 0.7071 |
| XGBoost | Multimodal | **0.9437** | **0.9147** | 0.8430 | **0.8773** |
| Transformer | Structured | 0.9194 | 0.8496 | 0.8655 | 0.8575 |
| Transformer | Notes | 0.8220 | 0.7591 | 0.7737 | 0.7663 |
| Transformer | Multimodal | 0.9209 | 0.8646 | 0.8625 | 0.8635 |

### 3.1 Bootstrap

#### XGBoost

Bootstrap analysis confirmed consistent performance across all three models for PR-AUC. The multimodal model achieved a median PR-AUC of 0.9436 with a 95% confidence interval of 0.9381 to 0.9494. For the Notes model, the median PR-AUC was 0.7204 (95% CI: 0.7047 – 0.7364), and for the structured model, it was 0.9374 (95% CI: 0.9306 – 0.9436). These values are presented in the upper part of Figure 2.

**Figure 2.**
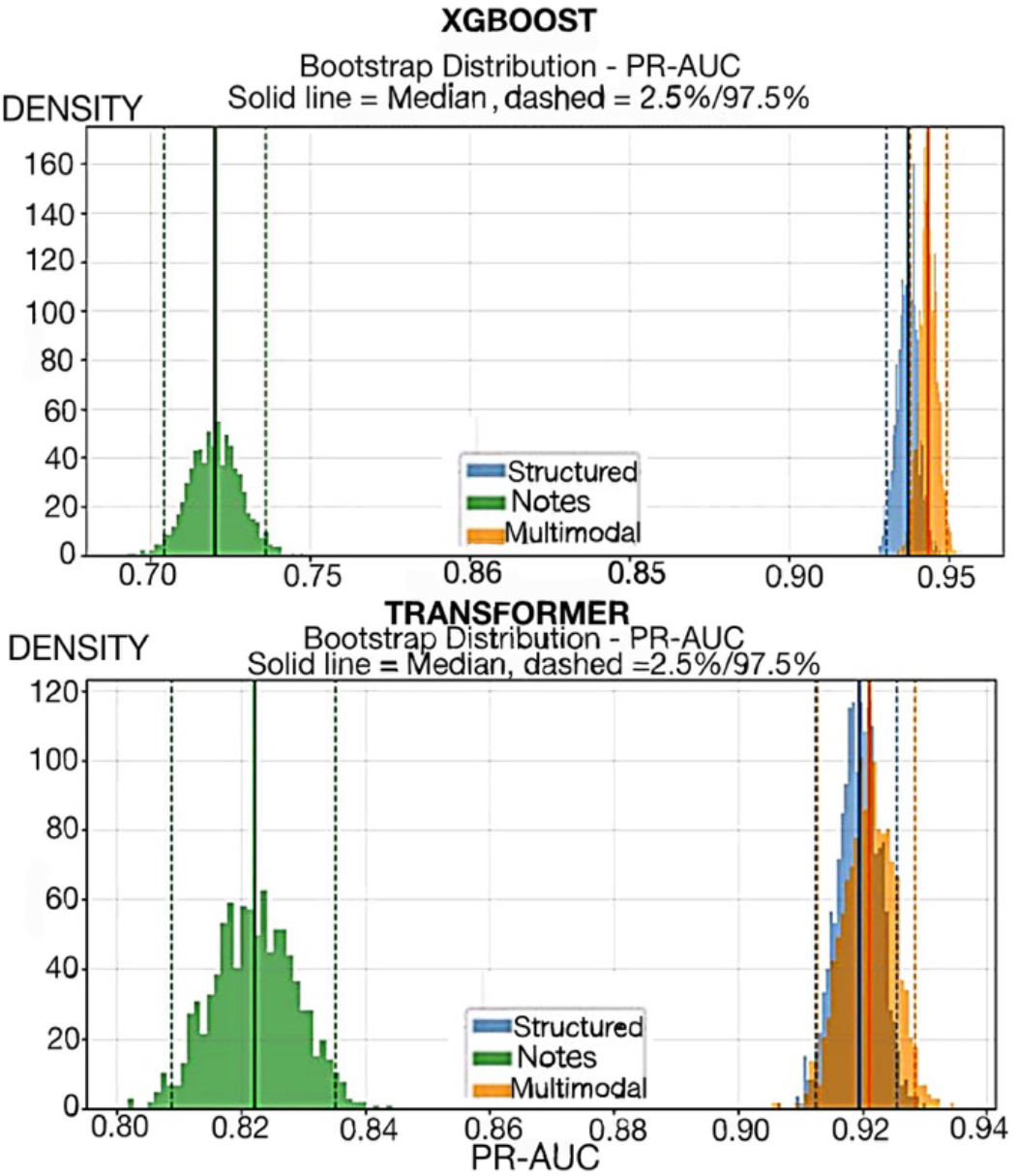
Bootstrap distributions of PR-AUC for XGBoost (top) and Transformer (bottom) models across structured(blue), notes(green), and multimodal(orange) configurations. Solid lines represent the median, while dashed lines indicate the 2.5% and 97.5% percentiles. The x-axis denotes PR-AUC values, and the y-axis represents density.

#### Transformer

The multimodal model achieved a median PR-AUC of 0.9208 with a 95% confidence interval of 0.9126 to 0.9287. For the Notes model, the median PR-AUC was 0.8221 (95% CI: 0.8086 – 0.8342), and for the structured model, it was 0.9195 (95% CI: 0.9119 – 0.9263). All values are shown at the bottom of Figure 2.

## 4 Discussion

Prior work has shown that XGBoost often outperforms DL models on structured clinical prediction tasks [8], with further studies confirming its competitiveness on tabular data [1, 9]. While multimodal Transformers have been proposed [13], they have not been directly compared with XGBoost under identical multimodal conditions for early HF prediction. Existing work remains focused on unimodal structured data. Multimodal models enable robust 48-hour prediction with

XGBoost achieving the best overall performance in line with previous findings on tabular clinical data [1] while Transformer perform better on clinical notes achieving higher precision and more reliable alerts whereas XGBoost achieves higher recall reducing missed HF cases. Compared with previously reported multimodal Transformer results (F1 of 0.49 and PR-AUC of 0.538) [13], our multimodal Transformer achieved substantially higher performance (F1 of 0.864 and PR-AUC of 0.921), highlighting the benefit of task-specific feature engineering and the integration of multimodal routine data. Bootstrap analysis confirmed model stability, showing only minor fluctuations across XGBoost variants, lower predictive value of clinical notes compared to structured data, and slightly improved performance in multimodal configurations. The Transformer showed greater stability across notes, resulting in more consistent predictions than XGBoost. The consistency of the results is crucial for integration into the dashboard, as it ensures stable risk assessments for patients. Several limitations remain. Validation in independent cohorts is necessary before clinical implementation. Since onset times were not always known, the model predicts early diagnosis rather than HF development. Additionally, the dashboard does not include note timestamps, reflecting a trade-off between documentation effort and usability that may become less relevant with future voice-based clinical documentation systems.

## 5 Conclusion

This study shows that a multimodal ML pipeline enables robust early HF diagnosis using MIMIC-IV data. Integrating multiple data modalities improved predictive performance over unimodal approaches, with multimodal XGBoost achieving the best overall metrics and the Transformer performing particularly well on notes. Both models were deployed in a dashboard to assist clinicians in early HF detection, potentially reducing hospitalizations and speeding diagnosis. While prior studies confirm XGBoost’s strength on structured clinical data [1, 9], our results extend this to a multimodal setting with unstructured notes, where Transformer provide complementary gains [13].

## Data Availability

All data utilized in the present work are available online at PhysioNet (https://physionet.org/content/mimiciv/3.1/, https://doi.org/10.13026/kpb9-mt58 and for the note database https://doi.org/10.13026/1n74-ne17, https://physionet.org/content/mimic-iv-note/2.2/). Access to the MIMIC-IV dataset is restricted to credentialed users who have completed the required training in human subjects research and signed a data use agreement

https://doi.org/10.13026/1n74-ne17

## Author Statement

Research funding: The author state no funding involved. Conflict of interest: Authors state no conflict of interest.

